# An integrated polygenic score stratifies risk of peripheral artery disease and adverse limb events in ancestrally diverse cohorts

**DOI:** 10.1101/2024.10.04.24314847

**Authors:** Alyssa M. Flores, Yunfeng Ruan, Anika Misra, Margaret S. Selvaraj, Tiffany R. Bellomo, Tetsushi Nakao, Kenneth Rosenfield, Matthew Eagleton, Whitney Hornsby, Aniruddh P. Patel, Pradeep Natarajan

## Abstract

**Background and Aims:** Peripheral artery disease (PAD) is a heritable atherosclerotic condition that is underdiagnosed and undertreated. With growing knowledge of the genetic basis for PAD and related risk factors, this study sought to construct a new polygenic score for PAD (GPS_PAD_).

**Methods:** GPS_PAD_ was constructed by integrating multi-ancestry summary statistics for PAD and related traits. GPS_PAD_ was trained in a UK Biobank dataset of 96,239 individuals and validated in a holdout UK Biobank dataset (N=304,294) and All of Us (AoU; N=237,173) and Mass General Brigham Biobank (MGBB, N=37,017).

**Results:** GPS_PAD_ was associated with an OR-per SD increase of 1.64 in the UK Biobank dataset (95% CI 1.60-1.68). Compared to previously published PAD polygenic scores, GPS_PAD_ was more strongly associated with PAD in AoU and MGBB, including enhanced transferability to non-European subgroups. GPS_PAD_ improved discrimination of incident PAD (ΔC-statistic 0.030) that was nearly equivalent to the additive performances of diabetes (ΔC-statistic 0.029) and smoking (ΔC-statistic 0.034). GPS_PAD_ was associated with reduced ankle-brachial index in the MGBB with the top 8% of individuals having a mean ABI ≤0.90 when assessed. Among individuals with prevalent PAD, GPS_PAD_ consistently identified individuals at high MALE-risk in the UK Biobank (HR 1.48; 95% CI 1.24-1.77), MGBB, (HR 1.34; 95% CI 1.12-1.60), and AoU (HR 1.33; 95% CI 1.12-1.58).

**Conclusions:** An integrated, multi-ancestry polygenic score for PAD predicts disease and adverse limb outcomes in three diverse cohorts. Incorporating polygenic risk into PAD care has the potential to guide screening and tailor management to prevent MALE.

## Introduction

Peripheral artery disease (PAD) is an atherosclerotic vascular condition that affects a global population of 230 million adults with high resource utilization owing to both systemic and limb ischemic events.^1,2^ While PAD shares risk factors with coronary artery disease (CAD), 32-54% of individuals presenting with PAD do not have clinically significant coronary or cerebrovascular disease.^3,4^ In addition, there are differences in the major etiologies of acute events in CAD and PAD. Ischemic CAD events most commonly result from plaque rupture, whereas the cause of acute limb ischemia is most commonly owing to embolism or *in situ* thrombosis, regardless of atherosclerosis extent.^5^ Indeed, the increasingly recognized role of thrombosis in PAD is also supported by the discovery of genetic variants in coagulation factors, including mutations in Factor V Leiden, which have been uniquely associated with PAD and not CAD.^6^

Clinical subsets of PAD include asymptomatic, claudication, and chronic-limb threatening ischemia (CLTI) resulting in tissue loss and major adverse limb events (MALE). MALE are a devastating complication and are often associated with critical illness, numerous resource-intensive attempts at revascularization, and prolonged hospital stays as part of the limb salvage effort. Despite its high morbidity, PAD is grossly underdiagnosed and has a lack of consensus on screening indications. For example, while European and American guidelines recommend consideration of screening based on age and risk factors, the United States Preventive Services Task Force (USPSTF) does not recommend screening regardless of risk factors.^7-9^ Conflicting recommendations have implications on care implementation, for example, in the U.S. where preventive service coverage is guided by USPSTF recommendations.^10^

In addition, there is no standard tool to predict complications before advanced disease develops. Such a tool would be useful as there are contemporary antithrombotic treatments targeted to prevent MALE but remain underutilized in PAD management.^11^

Polygenic risk scores (PRS) provide a quantitative metric for the inherited component of disease risk by integrating common genetic variants discovered from genome-wide association studies (GWAS) into a single instrument. As PAD has an estimated heritability of 20-50%^12,13^, incorporation of genetic factors offers an opportunity to optimize risk stratification. While there have been advances in PRS for CAD, there has been limited progress thus far in developments for PAD owing to largely European-based data, lack of validation in external datasets, and unclear transferability across diverse ancestry groups.^14-16^ Additionally, PRS utility for incident prediction of adverse PAD events has not been described.

To address these needs, in the present study, we develop a new genome-wide polygenic score for PAD (GPS_PAD_) that incorporates multi-ancestry GWAS data for PAD and related traits from over 2 million individuals (**Figure 1)**. We assess the performance of GPS_PAD_ in predicting PAD among individuals of diverse ancestry in a 304,294 internal validation cohort and two independent study populations comprising 259,627 individuals. Lastly, we apply GPS_PAD_ to predict MALE and identify individuals with clinically important increased risk.

**Figure 1.**
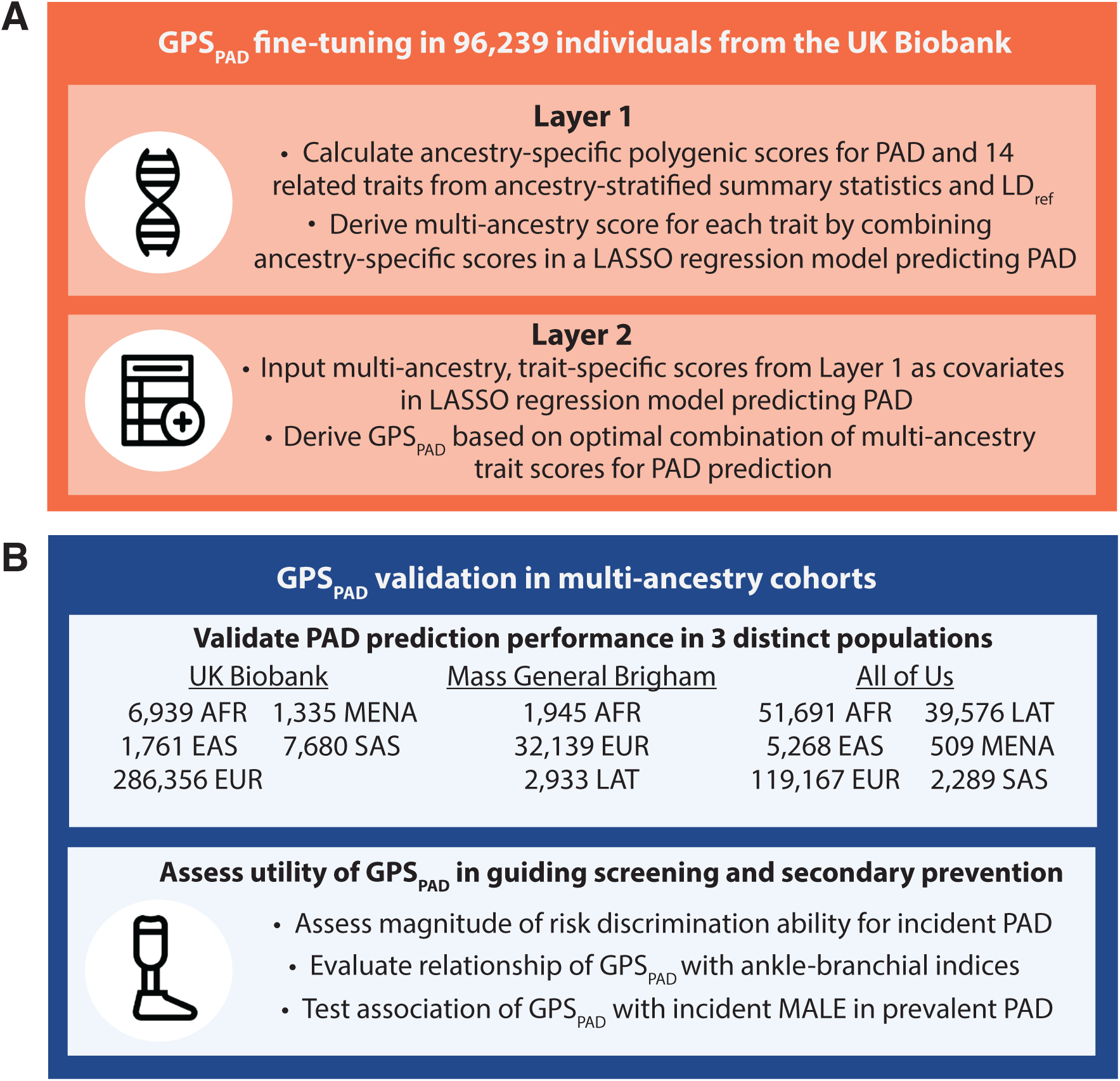
Schematic of GPS_PAD_ construction and study design. 1A. Overview of GPS_PAD_ development using GWAS summary statistics for PAD and 14 related traits stratified by African, East Asian, European, Latino, or South Asian ancestry. 1B. GPS_PAD_ was then evaluated in three validation datasets consisting of distinct, ancestrally diverse individuals, with outcomes including PAD (prevalent and incident), ankle-brachial indices, and MALE. Diagram designed with FreePik. AFR, African. EAS, East Asian. EUR, European. LAT, Latino. LD_ref,_ ancestry-matched LD reference panel. MENA, Middle Eastern/North African. SAS, South Asian.

## Methods

### Study populations

GPS_PAD_ was developed using data from the UK Biobank, a longitudinal cohort study of approximately 500,000 individuals from the United Kingdom aged 40-69 years.^17^ Participants were followed for outcomes based on International Classification of Disease 9^th^ and 10^th^ revisions (ICD-9/ICD-10) and Office of Population Censuses and Surveys versions 3 and 4 (OPCS-3/OPCS-4). Ancestry was based on self-identified ethnicity and country of origin with African, East Asian, European, Latino, Middle Eastern/North African (MENA), and South Asian categories (Supplemental Tables 1-2). Coding of comorbidities and lifestyle factors is detailed in the Supplemental Materials (Supplemental Table 3).

GPS_PAD_ performance was evaluated in two external datasets: Mass General Brigham Biobank (MGBB) and All of Us (AoU). The MGBB is a New England-healthcare based cohort that includes >145,000 patients treated at seven regional hospitals and clinics, including ∼56,000 have been genotyped.^18^ Baseline phenotypes are linked to the electronic health record (EHR) with available data from clinical notes, ICD-9/ICD-10, and Current Procedural Terminology (CPT) codes.

The AoU Research program is a United States-based cohort study that aims to recruit individuals who have been historically underrepresented in biomedical research.^19^ The program has recruited ∼400,000 individuals and includes health questionnaires, biospecimen collection, and longitudinal EHR data.

### Genetic data and quality control

In the UK Biobank, individuals were genotyped using UK BiLEVE Axiom Array or UK Biobank Axiom Array and centrally imputed to the 1000 Genomes (1000G) Panel, Haplotype Reference Consortium, or UK10K Panel.^17,20^ After performing quality control (Details Methods) and excluding individuals of Latino ancestry due to low population representation (N=11 PAD cases) and unreported/mixed ethnicity, 304,294 individuals were included for internal validation.

MGBB samples were genotyped on Illumina Multi-Ethnic Genotyping Array or Global Screening Array.^21^ Imputation was performed to the multi-ancestry TOPMED r2 reference panel. Given lack of detailed self-reported ethnicity or ancestry data, ancestry was genetically predicted using a K-nearest neighbor model trained with principal components (PCs) from the 1000G reference panels for European, African, Latino/Ad Mixed American, East Asian, and South Asian (Detailed Methods).^18^ We excluded individuals who were in prior discovery GWAS for CAD^22^ and did not map to a single genetic ancestry, leaving 37,017 individuals for external validation.

AoU participants were genotyped using the Illumina Global Diversity Array at AoU genome centers.^23^ Similar to the MGBB, there was a lack of detailed reported ancestry data, thus ancestry was assigned based on genetic similarity. AoU assigns categorical ancestries to African, Latino/Ad Mixed American, East Asian, South Asian, European, MENA, and Other based on a random forest classifier trained using gnomAD, Human Genome Diversity Project, and 1000G reference labels (Detailed Methods).^23^ After exclusion of individuals with missing/other ancestry, 237,173 individuals were included.

### Clinical endpoints

In the UK Biobank, PAD was defined based on self-reported history, ICD codes, OPCS codes for lower extremity revascularization or major amputation, and cilostazol prescription (Supplemental Table 4). In AoU, PAD was defined on self-report, occurrences of ≥2 ICD codes, or a single procedure code for revascularization, major amputation or supervised exercise therapy for PAD (Supplemental Table 5).^24^

In the MGBB, PAD was derived from phenotypes developed by the Mass General Brigham Research Patient Data Registry (RPDR) based on structured and unstructured clinical data from the EHR (Detailed Methods).^25^ ABI were extracted from imaging reports retrieved from the RPDR. We selected each individual’s minimum ABI and excluded values that were >1.4 or non-compressible.

MALE was defined as a surrogate of major amputation and acute limb ischemia based on diagnosis and procedure codes (Supplemental Tables 6-8). Revascularization included thrombectomy, thrombolysis, and emergency lower extremity bypass. In the MGBB, CPT codes for catheter-directed thrombolysis/thrombectomy required concomitant coding for aortogram or lower extremity angiography to ensure arterial intervention.

### GPS_PAD_ construction

PRS predictive performance is improved by the incorporation of data from increasingly diverse genetic ancestries and consideration of genetically correlated traits.^26-28^ To leverage the common mechanistic pathways of PAD, other atherosclerotic conditions, and PAD risk factors, GWAS results of PAD and 14 candidate traits were considered in GPS_PAD_ construction: PAD, CAD, ischemic stroke, glomerular filtration rate, diabetes mellitus, smoking, systolic blood pressure (SBP), diastolic blood pressure (DBP), low-density lipoprotein cholesterol (LDL-C), total cholesterol, high-density lipoprotein cholesterol (HDL-C), triglycerides, BMI, carotid plaque burden, and carotid intima-media thickness (Supplemental Table 9). After collection of GWAS summary statistics, GPS_PAD_ was trained using target data from 96,239 European individuals in the UK Biobank. GWAS results from the UK Biobank were not used to construct GPS_PAD_ such that data for score development and training were non-overlapping.

GPS_PAD_ was developed in a two-layer process (**Figure 1**).^26^ Layer 1 involved using ancestry-stratified GWAS data for each trait to calculate multi-ancestry polygenic scores that were optimized according to their PAD predictive performance. Separate scores were constructed using LDpred2, a widely used method that adjusts marginal single nucleotide polymorphism effect sizes for linkage disequilibrium (LD) patterns and selects a subset of variants with non-zero effects to calculate the polygenic score.^29^ Using LDpred2-auto, scores were calculated using GWAS results stratified by African, East Asian, European, Latino, and South Asian ancestry (R bigsnpr v.11.4). This resulted in 100 scores across all ancestries and traits. For each trait, the ancestry-specific scores were combined into the best-performing multi-ancestry score per trait using Least Absolute Shrinkage and Selection Operator (LASSO) regression in models predicting PAD (R glmnet v4.0-2). Feature selection was performed iteratively for all 15 traits in layer 1, yielding 15 multi-ancestry trait-specific scores with mixing weights detailed in Supplemental Table 9.

Layer 2 involved combining trait-specific scores from layer 1 to derive the integrated, multi-trait GPS_PAD_. Trait scores from Layer 1 were input into a LASSO regression model predicting PAD to construct the final GPS_PAD_. The final weight in GPS_PAD_ was derived by calculating the proportional weight from layer 1 in layer 2, normalized to 100%. Of the 15 candidate trait scores, 11 traits contributed to GPS_PAD_ with mixing weights in Supplemental Table 10. GPS_PAD_ was then calculated in the validation UK Biobank dataset. See Detailed Methods for details on GPS_PAD_ construction.

### GPS_PAD_ validation and benchmarking

GPS_PAD_ was calculated in the MGBB and AoU for external validation. GPS_PAD_ performance was compared with previously published PRS in the Polygenic Score Catalog^30^ and a recently reported multi-ancestry polygenic score for CAD (GPS_CAD_).^26^ We previously developed GPS_CAD_ utilizing a similar two-layer framework for CAD as in the present study with slight modifications including stepwise regression for feature selection.^26^ From the Polygenic Score Catalog, PRS_PLR_ is a single-trait score for PAD developed using LASSO regression and individual-level European data from the UK Biobank.^15^ PRS_LDpred2_ is another single-trait score for PAD trained using UK Biobank data, but was calculated with LDpred2-auto.^15^

We also calculated the lifetime risk of PAD in individuals in the UK Biobank based on clinical variables using the Johns Hopkins University PAD risk tool (http://ckdpcrisk.org/padrisk, Detailed Methods).^31^

### Statistical analysis

The unadjusted rate of PAD was calculated across percentiles of GPS_PAD_. Model calibration was assessed with the Hosmer–Lemeshow test (R ResourceSelection v0.3-6) and by comparing the observed and predicted prevalence across percentiles calculated using a logistic regression model with only GPS_PAD_ as a predictor. We estimated PAD risk in the extremes of the GPS_PAD_ distribution using logistic regression models. We also derived the proportion of the population with a given magnitude of risk by calculating the odds ratio (OR) of varying extremes of GPS_PAD_ percentiles compared to the middle quintile group (40-59%).

The association of GPS_PAD_ and other scores with all PAD (incident and prevalent cases) were assessed using logistic regression. Performance metrics included OR, area under the receiver operator characteristic curve (AUC, R pROC v1.17.0.1), and phenotypic variance explained (Nagelkerke-R^2^). Incident event analyses were performed using Cox proportional hazards models with metrics including hazard ratio (HR) and C-statistic (R survival v3.5-7). MALE analyses were restricted to individuals with prevalent PAD diagnoses. Time-to-MALE curves were estimated using the Kaplan-Meier method, standardized to mean age and gender (R survminer v0.4.9). Linear regression was used to evaluate the relationship between polygenic scores and minimum ABI.

Logistic, linear, and Cox models including polygenic scores were adjusted for age, sex, genotyping array, and the first ten PCs. Cox models were used to estimate the 10-year incidence of PAD across lifestyle and genetic risk groups, standardized to the mean of covariates in each group. In the UK Biobank, we performed sensitivity analyses to test the strength of GPS_PAD_ associations with PAD and incident MALE after additionally accounting for clinical variables (current smoking, hypertension, diabetes, hyperlipidemia, and chronic kidney disease); and continuous variables for individuals with available physical and laboratory data (SBP, DBP, hemoglobin A1c, HDL-C, estimated untreated LDL-C^32^, GFR, and anti-hypertensive use).

GPS_PAD_ and other continuous variables were scaled to a mean of zero and one standard deviation (SD) such that effect sizes indicate OR- or HR-per SD. Statistical significance was defined as *P*<0.05 or 95% confidence interval (CI) that excluded the null value. Statistical analyses were performed using R-4.1.0.

## Results

Given that complex diseases such as PAD share genetic backgrounds with related traits, we leveraged this genetic correlation to more fully capture human genetic architecture and construct GPS_PAD_.^6,33,34^ GPS_PAD_ included 603,595 variants across 11 traits and 5 ancestries, including PAD, smoking, CAD, ischemic stroke, diabetes, SBP, LDL-C, glomerular filtration rate, BMI, carotid IMT, and total cholesterol. While GPS_PAD_ was predominantly derived from European discovery data, the score incorporated genetic variation discovered in African, East Asian, South Asian, and Latino populations (Supplemental Table 10). Of the non-European populations, African GWAS discovery data contributed the most to GPS_PAD_.

### Association of GPS_PAD_ with PAD risk

Within the UK Biobank, GPS_PAD_ was associated with a OR 1.77 (95% CI: 1.70-1.86) for PAD in the European training sample (Supplemental Table 12). The effect size was mildly attenuated in the multiethnic validation cohort including 304,294 individuals, but remained strongly associated with PAD (OR 1.63; 95% CI 1.60-1.68). The holdout validation cohort included 164,108 females (53.9%) and 286,356 participants of European (94%), 7,680 South Asian (2.5%), 6,939 African (2.3%), 1,761 East Asian (0.6%), and 1,558 (0.51%) MENA ancestry. There was a decrement, yet persistently significant association with adjustment for clinical risk factors (OR 1.35; 95% CI 1.31-1.39) and in a model adjusted for continuous covariates (OR 1.37; 95% CI: 1.33-1.41).

We found significant differences in PAD rates across the GPS_PAD_ percentile distribution, ranging from 0.78% in the bottom percentile to 7.91% in the top percentile (**Figure 2A**, Supplemental Table 13). Predicted PAD prevalence was overall consistent with observed prevalence, excluding the ≥98% percentile where there was slight risk underestimation by GPS_PAD_ alone driven by a sharp increase in the observed PAD prevalence (Supplemental Figure 1). GPS_PAD_ stratified individuals in low and high-risk groups with up to 4.71-fold increased PAD risk in the top 1% of GPS_PAD_ (OR 4.71, 95% CI 4.07-5.42) compared to the middle quintile (**Figure 2B**).

**Figure 2:**
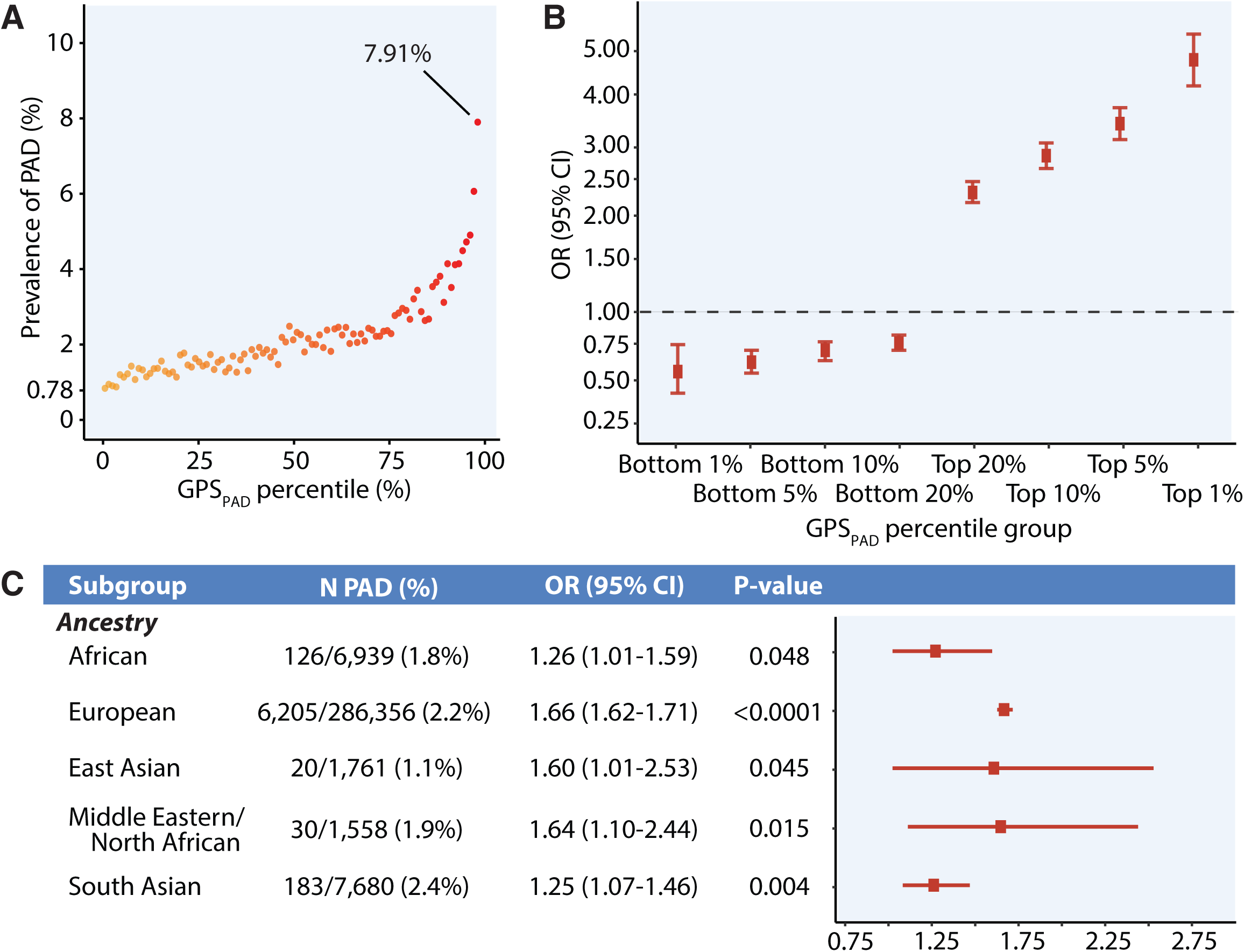
GPS_PAD_ is an integrated polygenic score that stratifies PAD risk in diverse ancestries. 2A. Prevalence of PAD across GPS_PAD_ percentiles in the UK Biobank validation dataset (N = 304,294). 2B. Estimated PAD risk in the top and bottom of GPS_PAD_ percentile distributions relative to the middle quintile (40-59%), as assessed in logistic regression models adjusted for age, sex, genotyping array, and ten principal components of ancestry. Results are shown on log-axis. 2C. Risk of PAD associated with GPS_PAD_ in ancestral subgroups in the UK Biobank. Results were calculated from a logistic regression model with age, sex, genotyping array, and ten principal components as covariates.

Compared to previously published scores, GPS_PAD_ resulted in an improvement in effect estimate, discrimination, and proportion of phenotypic variance explained compared to PRS_PLR ._ In a model adjusted for age, sex, genotyping array, and the first ten PCs of ancestry, GPS_PAD_ had an OR of 1.63, AUC of 0.74, and Nagelkerke-R^2^ of 0.087, while PRS_PLR_ had an OR of 1.20, AUC 0.71, and Nagelkerke-R^2^ of 0.064. PRS_LDpred2_ achieved greater performance in the UK Biobank than GPS_PAD_ with OR 2.46 and nearly equivalent discriminative ability with AUC 0.75, however Nagelkerke-R^2^ and effect sizes in the extremes of PRS_LDpred2_ percentiles strongly suggested overfitting. For example, PRS_LDpred2_ Nagelkerke-R^2^ was 0.17 in the UK Biobank validation cohort and OR for the top 1% PRS_LDpred2_ was 71.9 (95% CI 64.9-79.7).

GPS_PAD_ identified individuals with distinct risk compared to an integrated polygenic score for CAD (GPS_CAD_).^26^ In the UK Biobank, GPS_PAD_ was more strongly associated with PAD (OR 1.63; 95% CI 1.60-1.68) than GPS_CAD_ (OR 1.48; 95% CI 1.44-1.52) and identified a greater number of high-risk individuals (Supplemental Table 13). For example, the proportion of the UK Biobank population at 3-fold and 4-fold greater odds for PAD was 5.7% (N=17,345) and 1.90% (N=5,782) when classified by GPS_PAD_, respectively. There was a comparatively small fraction of individuals at similar PAD risk according to GPS_CAD_ with only 1.90% (N=5,782) at 3-fold greater risk and 0.20% (N=609) at 4-fold greater risk.

We then compared polygenic score performance in ancestral subgroups in the UK Biobank (**Figure 2C**). Across each ancestral population, GPS_PAD_ predicted disease risk in African, European, East Asian, MENA, and South Asian subgroups with effect estimates of 1.26, 1.66, 1.60, 1.64, and 1.25 respectively. While PRS_LDpred2_ and PRS_PLR_ predicted PAD risk in European individuals, both were not associated with PAD in non-European subgroups (Supplemental Table 14). In clinical subgroups, GPS_PAD_ performance was fairly stable, with OR ranges of 1.33-1.72 depending on demographics and the presence of risk factors (Supplemental Figure 2).

### Validation of GPS_PAD_ in distinct external cohorts

GPS_PAD_ was externally validated in two independent multi-ancestry cohorts and showed improved cross-population predictive performance relative to previously published scores. GPS_PAD_ was significantly associated with an increased risk for PAD (OR 1.26; 95% CI 1.18-1.37, **Figure 3A**) in the MGBB, which included 5.3% African (N=1,945), 87% European (N=32,139), and 7.9% Latino individuals (N=2,933). GPS_PAD_ performed similarly in AoU (OR 1.21; 95% CI 1.19-1.23), which embodied 22% African (N=51,691), 2.2% East Asian (N=5,268), 50% European (N=119,167), 17% Latino (N=39,576), 0.21% MENA (N=509), and 0.97% South Asian individuals (N=2,289). Of note, while PRS_LDpred2_ outperformed GPS_PAD_ in the UK Biobank where it was derived, PRS_LDpred2_ was not significantly associated with PAD risk in the MGBB (OR 1.02; 95% CI 0.95-1.08; *P*=0.06) and was more weakly associated with PAD than GPS_PAD_ in AoU (OR 1.10; 95% CI 1.08-1.12). GPS_PAD_ consistently had stronger effect sizes than PRS_PLR_ in each external cohort (**Figure 3A**).

**Figure 3.**
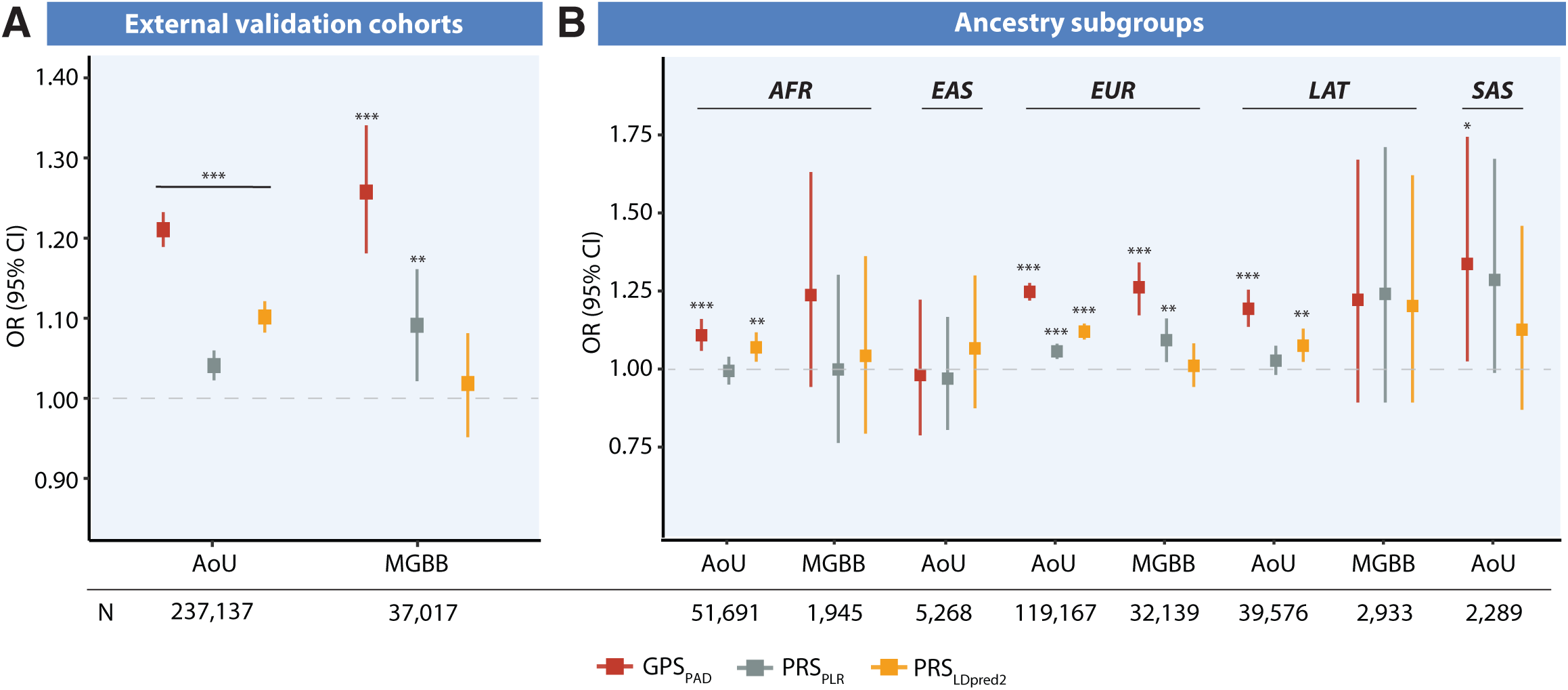
GPS_PAD_ shows improved transferability to external cohorts. 4A. OR per-SD change of polygenic scores in external validation cohorts, AoU (N = 237,173) and MGBB (N = 37,017). 4B. Predictive performance for each score is also shown in ancestry subgroups: African individuals in AoU (N = 51,691) and MGBB (N = 1,945); East Asians in AoU (N = 5,268), European individuals in AoU (N = 119,167) and MGBB (N = 32,139); Latino individuals in AoU (N = 39,576) and MGBB (N = 2,933); and South Asian individuals in AoU (N = 2,289). OR were calculated from a logistic regression model adjusted for age, sex, genotyping array, and the first ten principal components. Asterisks represent P-value for associations, *=P<0.05. **=P<0.01. ***P<0.0001. EAS, East Asian. EUR, European. LAT, Latino. SAS, South Asian.

When stratifying by ancestry, GPS_PAD_ demonstrated improved transferability in most subgroups (Supplemental Table 14). Among African (N=51,691), European (N=119,167), Latino (N=39,576), and South Asian (N=2,289) individuals in AoU, GPS_PAD_ was the most strongly predictive of PAD compared to previously published scores (**Figure 3B**). GPS_PAD_ was also transferable to Europeans in the MGBB with a larger effect size (OR 1.26; 95% CI 1.17-1.34) than PRS_PLR_ (OR 1.09; 95% CI 1.02-1.16) and PRS_LDpred2,_ which was not associated with disease risk (OR 1.01; 95% CI 0.94-1.08). No polygenic scores were associated with PAD in AoU East Asians, AoU MENA, and non-European subgroups in the MGBB (Supplemental Figure 3).

### Modeling incident PAD according to polygenic risk and clinical risk factors

We then analyzed 301,932 participants of the UK Biobank who were free of PAD at study enrollment. Over a median 12.1 years follow-up (interquartile range (IQR) 10.6-13.5 years), incident PAD was observed in 4,202 individuals (1.39%). Within the UK Biobank, the baseline model of age, sex, and ten PCs of ancestry had a C-statistic of 0.731 (**Figure 4A**). The addition of GPS_PAD_ resulted in improvement in discrimination to 0.761, which was greater than the improvement of most risk factors also in isolation, and was approximately equivalent to the additive benefit of diabetes (C-statistic 0.760) and smoking (C-statistic 0.765). The greatest improvement in discrimination was observed with the baseline model, clinical variables, and GPS_PAD_ (C-statistic 0.087).

**Figure 4.**
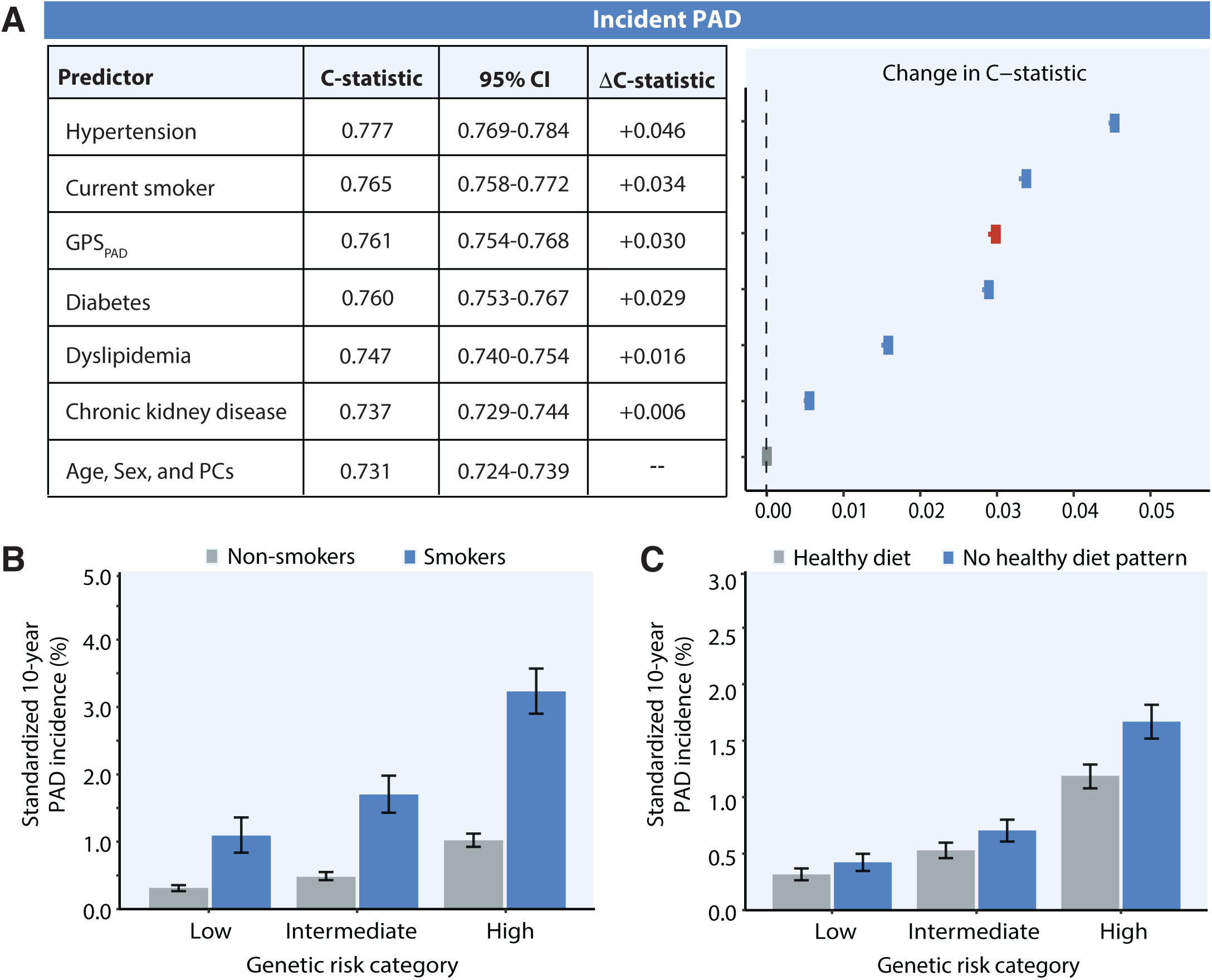
Performance of GPS_PAD_ in predicting incident PAD in the UK Biobank. 4A. Changes in C-statistic of GPS_PAD_ (red point) and clinical risk factors (blue points) relative to the baseline model including age, sex, and the first ten principal components of ancestry (gray point), as calculated from Cox proportional hazards models in the UK Biobank holdout validation dataset. Lines represent confidence intervals of the C-statistic change. 4B-C. 10-year rates of PAD according to lifestyle and genetic categories, showing the influence of smoking (4B) and dietary patterns (4C). Shown are standardized cumulative rates of incident PAD standardized to the population averages for each covariate in the Cox models.

Compared to the individual performance of other scores, GPS_PAD_ resulted in greater benefit in discrimination of incident PAD than PRS_PLR_ (C-statistic 0.735) and the lifetime PAD risk score^31^ (C-statistic 0.675). PRS_LDpred2_ achieved a larger improvement in C-statistic (C-statistic 0.774) from the baseline model compared to GPS_PAD_, but the HR was drastically large in the UK Biobank where it was wholly derived, and consistent with overfitting (HR 199; 95% CI 172-231), as observed in cross-sectional analyses.

In the baseline model, GPS_PAD_ HR for incident PAD was 1.66 (95% CI 1.61-1.71) and HR 1.37 (95% CI 1.30-1.46) in a multivariable model adjusted for clinical risk factors. When examining the interaction between GPS_PAD_ and clinical variables, we observed a weak interaction with levels of LDL-C (GPS_PAD_*LDL-C HR 1.05; 95% CI 1.01-1.08; *P_interaction_*=0.005) and hemoglobin A1c (GPS_PAD_*hemoglobin A1c HR 1.02; 95% CI 1.00-1.03; *P_interaction_*=0.01). Lifestyle factors influenced PAD risk across each stratum of genetic risk (Supplemental Table 15). For the high genetic risk group, the standardized 10-year incidence of PAD of 3.17% for smokers and decreased to 1.05% for those who refrained from smoking (**Figure 4A)**. There were similar patterns of an offset of genetic risk by modifiable lifestyle factors according to dietary patterns (**Figure 4B**).

### Integration of polygenic risk with clinical guidelines

We then assessed whether GPS_PAD_ could identify individuals at equivalent risk for incident disease as the high-risk groups that current guidelines recommend screening for PAD.^8^ We first quantified the 10-year incidence of PAD among subgroups of individuals proposed to be candidates for ABI screening according to the AHA/ACC Guidelines.^8^ Individuals above 65y had a 10-year incidence of 2.28% (95% CI 2.16-2.41), individuals who were age 50-64y with one risk factor for atherosclerosis had an incidence of 1.23% (95% CI 1.17-1.30%), individuals <50y with diabetes and 1 additional risk factor had an incidence of 2.90% (95% CI 1.62-3.21%), and those with known atherosclerosis in another vascular bed had the greatest incidence of 5.95% (95% CI 5.48-6.42). As a comparison, incidence rates ranged from 2.81-4.40% in the top 1-5% GPS_PAD_ compared to the middle quintile distribution (Supplemental Table 16). We also looked at age-based combinations of each individual risk factor and compared event rates to subgroups with the combination of risk factors and high polygenic risk.

This analysis revealed that the combination of high GPS_PAD_ with one atherosclerosis risk factor identified more individuals who subsequently developed PAD than many of the subgroups defined by the combination of similar risk factors with age (**Figure 5A)**. For example, 7.96% of individuals in the top 5% GPS_PAD_ and diabetes developed PAD, as compared to a 4.02% incident rate among individuals aged 50-64y with diabetes.

**Figure 5.**
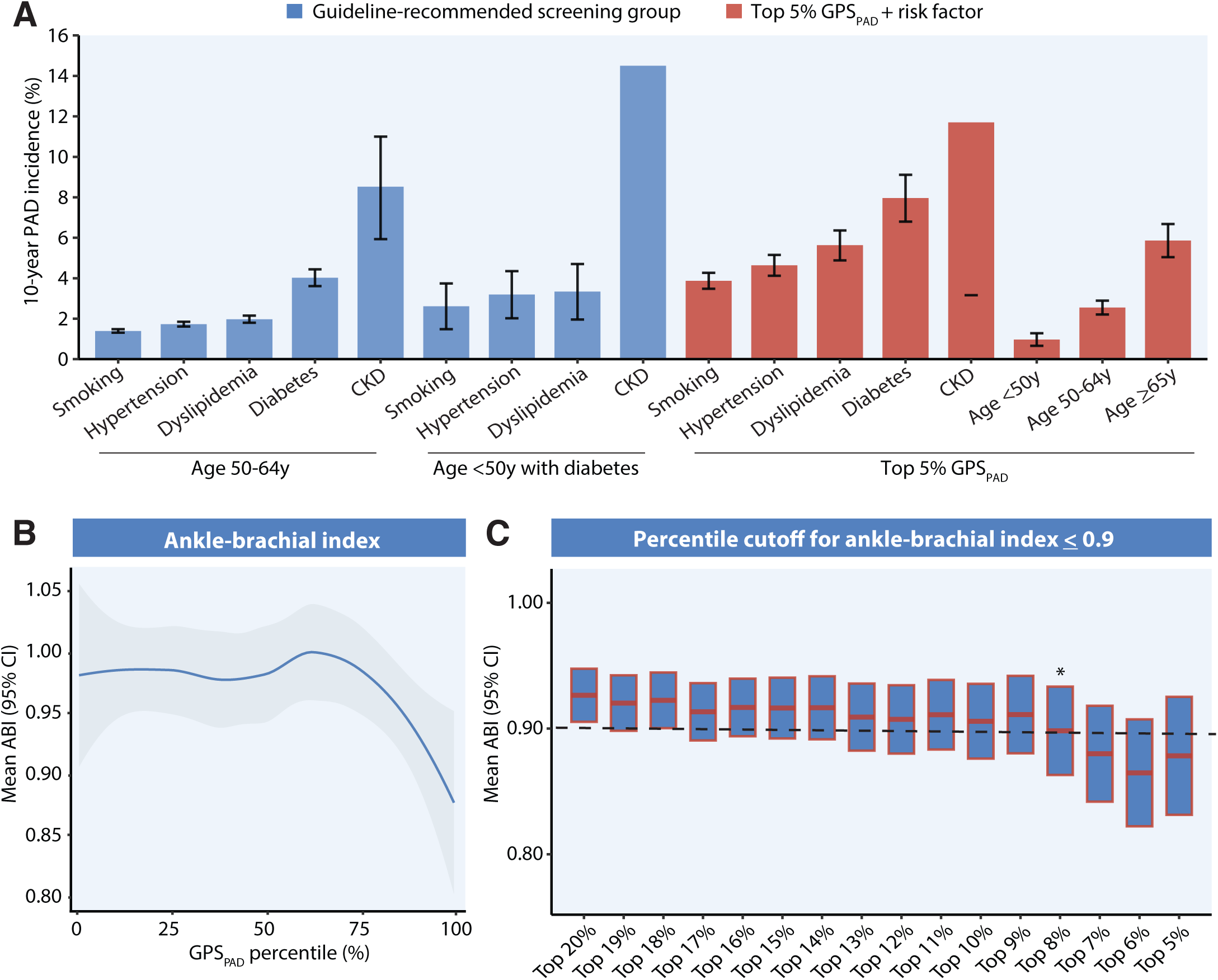
Incorporation of GPS_PAD_ into screening criteria and PAD diagnosis. 5A. 10-year incidence of first-occurring PAD events from prospective analyses of individuals without prior PAD in the UK Biobank. Cox proportional hazards regression models were used to calculate the 10-year event risk among groups for which current clinical guidelines recommend ABI screening^8^ (blue) and individuals in the top 5% GPS_PAD_ with individual risk factors (red). Error bars represent 95% CI and are not completely shown for groups in the age<50y with diabetes + CKD or top 5% GPS_PAD_ + CKD, but can be found in Supplemental Table 16. 5B. Loess regression plot of GPS_PAD_ and minimum ABI among a subset of individuals in the Mass General Brigham Biobank (N = 763). 5C. Mean ABI for individuals in the top percentiles of GPS_PAD._ * indicates percentile group with mean ABI≤0.9. AAA, abdominal aortic aneurysm. CKD, chronic kidney disease.

Similarly, there was a relative increase in PAD cases among individuals in the top 5% GPS_PAD_ with dyslipidemia (5.63%; 95% CI 4.88-6.36) compared to individuals 50-64y with dyslipidemia (1.97%; 95% CI 1.80-2.15). The addition of the top 5% GPS_PAD_ also increased the proportion of additional cases of PAD when combined with other atherosclerotic conditions such as CAD and carotid stenosis compared to consideration of the conditions alone (Supplemental Table 16).

### Predicting ankle-brachial indices based on polygenic risk

In addition to predicting binary PAD presence, we evaluated whether GPS_PAD_ was associated with minimum ABI for a subset of individuals in the MGBB (N=883, Supplemental Figure 4). The minimum ABI was first used to validate the PAD phenotype in the MGBB. PAD cases indeed had a lower mean ABI compared to controls (0.78 vs. 1.10; P<2x10^-16^, Supplemental Figure 5). After adjusting for age, sex, and the first ten PCs, GPS_PAD_ was significantly associated with lower ABI (**Figure 5B**), while previously published scores were not (all *P*>0.1, Supplemental Table 17). Each SD increase in GPS_PAD_ was associated with an -0.088 reduction in ABI (95% CI -0.161 to -0.015; *P*=0.019). Based on the percentile distribution in the MGBB, the group in the top 8% GPS_PAD_ had a mean ABI≤0.90 diagnostic for PAD (95% CI 0.83-0.97, **Figure 5C)**.^35^

### Association of GPS_PAD_ with incident major adverse limb events

Lastly, we assessed whether polygenic background could identify individuals at increased risk for MALE after PAD diagnosis. In external validation cohorts, GPS_PAD_ was significantly associated with incident MALE and provided improved risk stratification than previously published scores. Individuals with high polygenic risk (top 20% of GPS_PAD_) in the UK Biobank had a 75% higher relative risk of incident MALE (HR 1.75; 95% CI 1.18-2.57; *P*=0.005) over a median of 12.9 years (IQR 11.7-14.1) compared to the remainder of the PAD population. The top 20% GPS_PAD_ also had an increased risk of MALE in the MGBB (HR 1.56; 95% CI 1.06-2.30; *P*=0.02) during 5.19 years of follow-up (IQR 2.63-7.75) and AoU cohort (HR 1.51; 95% CI 1.03-2.24; *P*=0.03) over 2.32 years of follow-up (IQR 0.35-4.29, **Figure 6A**).

**Figure 6.**
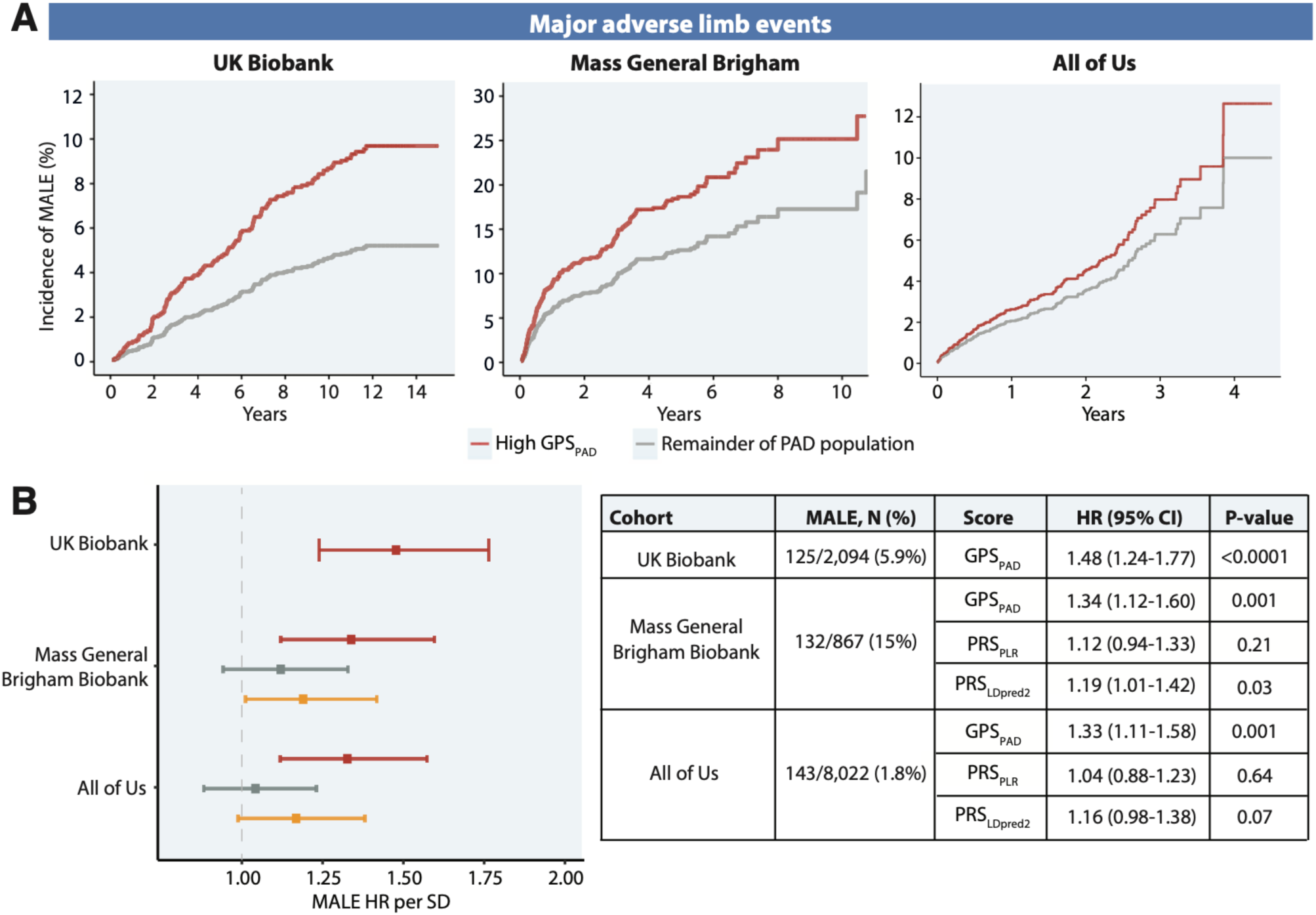
GPS_PAD_ stratifies risk for incident major adverse limb events among individuals with PAD. 6A. Kaplan-Meier curves of MALE according to high GPS_PAD_ compared to the remainder of the PAD population, standardized to the mean age and sex of each cohort. 6B. Risk associated with polygenic scores for incident MALE among individuals with diagnosed PAD in the UK Biobank (N = 2,094), Mass General Brigham Biobank (N = 867), and All of Us (N = 8,022). HRs were calculated from adjusted Cox regression models including age, sex, genotyping array, the first ten principal components of ancestry as covariates.

In the overall PAD cohort in the UK Biobank, GPS_PAD_ was significantly associated with MALE (HR 1.48; 95**%** CI 1.24-1.77; **Figure 6B**), which was diminished but remained associated after multivariable adjustment (HR 1.30; 95% CI 1.07-1.59; *P*=0.009).

Associations of GPS_PAD_ with incident MALE remained significant in the prevalent PAD cohorts of the MGBB (HR 1.34; 95% CI 1.12-1.60) and AoU (HR 1.33; 95% CI 1.11-1.58; **Figure 6B**), while other scores demonstrated weaker or non-significant risk estimates.

## Discussion

Using ancestry-stratified GWAS results from multiple populations, we developed a polygenic score that aggregates the effects of variants in PAD and related traits across five ancestries. GPS_PAD_ shows strong disease prediction and is transferable to external cohorts with ancestrally diverse populations, demonstrating associations with lower ABI and greater effect sizes for PAD association in non-European populations.

This new score also identifies more high-risk individuals than a polygenic score for CAD, highlighting the differences in genetic risk for these atherosclerotic conditions.^28^ Moreover, among individuals with diagnosed PAD, high GPS_PAD_ predicts future risk of MALE, defined as a surrogate of major amputation and acute limb ischemia.

Prior polygenic scores for PAD were solely derived from European data or employed GWAS data from a single population and were not validated in external cohorts.^14-16^ Our computational framework considered variants discovered from >2 million ancestrally diverse individuals and leveraged the pleiotropy between PAD and related risk factors to enhance genetic risk prediction.^26^ Such approaches that incorporate genome-wide genetic correlation increase the power of polygenic association analyses and improve prediction accuracy.^36,37^ GPS_PAD_ was indeed the most strongly predictive of PAD risk in external validation cohorts. Unlike previously published polygenic scores, GPS_PAD_ was able to predict PAD risk in Latino individuals in All of Us and both African and South Asian individuals in two distinct cohorts (All of Us and the UK Biobank).

In prospective analysis in the UK Biobank, GPS_PAD_ was associated with incident PAD with similar effect sizes as cross-sectional analysis. The inclusion of GPS_PAD_ resulted in a comparable benefit in discriminative capacity as those afforded by smoking and diabetes.^3^ We also note a significant interaction between GPS_PAD_ and LDL-C or hemoglobin A1c in models predicting incident PAD. This suggests that individuals with higher GPS_PAD_ may have a greater benefit from dietary changes, statin therapy, or glycemic control. Indeed, post-hoc analyses of statin therapy trials demonstrate that statins confer a greater cardiovascular risk reduction in those with high genetic risk compared to the general population.^38^

Current European guidelines more broadly recommend consideration of screening based on age and cardiovascular risk factors, while American cardiovascular society guidelines recommend screening for PAD with ABI in four groups: age≥65 years, age 50-64 years with atherosclerotic risk factors, age <50 years with diabetes and 1 risk factor, and history of atherosclerosis in other vascular beds.^8,9^ Addition of the GPS_PAD_ has the potential to further streamline these screening recommendations and identify higher risk populations. When evaluating the combination of polygenic risk and clinical risk assessment, incorporation of the top 5% polygenic risk with conventional risk factors identified more individuals who went on to develop PAD compared to the combination of risk factors with age. These results suggest the top 5% genetic risk and presence of risk factors could be considered a criteria for screening with ABIs to establish PAD diagnosis. Indeed, when leveraging ABIs, our analysis revealed an ABI diagnostic of PAD associated with high GPS_PAD_ in the MGBB, although replication in other cohorts is needed.

Secondary prevention efforts in PAD focus on limb salvage. MALE are associated with poor prognosis for PAD patients and represent index events for subsequent hospitalizations, reinterventions, major adverse cardiac events, and even mortality.^39^ Once acute limb ischemia occurs, numerous returns to the operating room for revascularization with combined exposure to anticoagulation and thrombolytic therapy and subsequent reperfusion is a major physiologic stress in an already comorbid patient population. Staging systems such as the Wound, Ischemia, and foot Infection are used to determine amputation risk in the CLTI population, but there are no current vetted clinical tools for predicting MALE early in the disease course.^3^ Our analysis found that GPS_PAD_ stratified MALE risk specifically in the PAD population and remained consistently predictive of incident MALE in the UK Biobank, Mass General Brigham Biobank, and All of Us.

These findings have several implications. First, an individual’s genetic risk in combination with their cardiovascular risk factor profile may be employed as a criteria for consideration of screening with non-invasive vascular testing. PAD is an underdiagnosed and undertreated condition with guideline-directed therapies that lag that of CAD.^8^ Incorporation of polygenic risk may be used as an adjunct to clinical risk assessment to identify a greater proportion of high-risk individuals for medical optimization. In addition, as with studies of polygenic risk and CAD^38,40-42^, there may be early benefits for targeted intensive lifestyle and risk factor optimization in PAD with high genetic risk. However, there is need for further testing the effect of polygenic scores on hard cardiovascular outcomes in prospective studies, particularly among younger age groups. Given that high GPS_PAD_ was strongly associated with MALE in three distinct PAD cohorts, there is promise for implementation of polygenic scores into secondary prevention measures for vascular practitioners, such as in frequency of surveillance or guiding more aggressive or prolonged antithrombotic therapies shown to reduce MALE incidence.^11,39^

Our study has several limitations. First, GPS_PAD_ was trained using data from largely European individuals due to limitations in sample size of other ancestries. Nevertheless, we demonstrated evidence of portability to non-European groups, not previously achieved by prior PAD polygenic scores. Second, this study was restricted to individuals with genetic similarities to single ancestries and had more limited cases of non-European groups, which could have limited power to detect associations especially among the East Asian and MENA groups. Race alone was importantly not used to infer ancestry, however such broad ancestral categorizations can mask heterogeneity in polygenic score performance in some groups. Tailoring of polygenic scores to non-European target data, continued recruitment of underrepresented populations in biobanks, and development of methods to account for admixture should be a priority to improve polygenic score performance. Lastly, PAD and MALE phenotyping were based on diagnosis and procedures codes from the EHR. There could have been variation in the quality of definitions between cohorts that influenced analyses towards null associations. However, we used non-invasive studies in the MGBB as another surrogate for PAD and observed similar associations of polygenic risk with binary PAD classifications in both the MGBB and AoU, where the latter is comprised of several U.S. healthcare institutions.

## Ethics Approval

Individuals in the UK Biobank, All of Us, and Mass General Brigham Biobank underwent signed consent for genetic sequencing, storage of biological specimens, and access to electronic health record data. This research was conducted using the UK Biobank resource under application number 7089. Secondary analyses of the UK Biobank, MGBB, and AoU was approved by the Massachusetts General Hospital Institutional Review Board.

## Data Availability

Individual-level data from the UK Biobank and All of Us are available upon request from researchers to each organization. This study used Controlled Tier data from All of Us which is available to authorized users on the All of Us Researcher Workbench. GPS_PAD_ constructed in this paper will be made available in the Polygenic Score Catalog following publication. Polygenic scores used for comparison of GPSPAD performance are available in the Polygenic Score Catalog through accession ID PGS001843 and PGS002055.

## Acknowledgments

We thank the participants and investigators in the UK Biobank, All of Us, and Mass General Brigham Biobank for their contributions to this study.

## Abbreviations

AoU: All of Us
ABI: Ankle-brachial index
BMI: Body mass index
CAD: Coronary artery disease
CPT: Current Procedural Terminology
DBP: Diastolic blood pressure
EHR: Electronic health record
GWAS: Genome-wide association study
GFR: Glomerular filtration rate
HDL-C: High-density lipoprotein cholesterol
ICD: International Classification of Disease
LASSO: Least Absolute Shrinkage and Selection Operator
LDL-C: Low-density lipoprotein cholesterol
MALE: Major adverse limb events
MGBB: Mass General Brigham Biobank
OPCS: Office of Population Censuses and Surveys
PAD: Peripheral artery disease
PRS: Polygenic risk scores
PCs: Principal components
RPDR: Research Patient Data Registry
SBP: Systolic blood pressure
USPSTF: United States Preventive Services Task Force

## References

1. Song P, Rudan D, Zhu Y, et al. Global, regional, and national prevalence and risk factors for peripheral artery disease in 2015: an updated systematic review and analysis. Lancet Glob Health 2019;7:e1020–e1030. doi: 10.1016/S2214-109X(19)30255-4

2. Mahoney EM, Wang K, Keo HH, et al. Vascular hospitalization rates and costs in patients with peripheral artery disease in the United States. Circ Cardiovasc Qual Outcomes 2010;3:642–651. doi: 10.1161/CIRCOUTCOMES.109.930735

3. Aday AW, Matsushita K. Epidemiology of Peripheral Artery Disease and Polyvascular Disease. Circ Res 2021;128:1818–1832. doi: 10.1161/CIRCRESAHA.121.318535

4. Hackler EL, 3rd, Hamburg NM, White Solaru KT. Racial and Ethnic Disparities in Peripheral Artery Disease. Circ Res 2021;128:1913–1926. doi: 10.1161/CIRCRESAHA.121.318243

5. Narula N, Olin JW, Narula N. Pathologic Disparities Between Peripheral Artery Disease and Coronary Artery Disease. Arterioscler Thromb Vasc Biol 2020;40:1982–1989. doi: 10.1161/ATVBAHA.119.312864

6. Klarin D, Lynch J, Aragam K, et al. Genome-wide association study of peripheral artery disease in the Million Veteran Program. Nat Med 2019;25:1274–1279. doi: 10.1038/s41591-019-0492-5

7. Mills J, Duffy M. Screening for Peripheral Artery Disease and Cardiovascular Disease Risk Assessment with the Ankle-Brachial Index. Am Fam Physician 2018;98:754–755. doi:

8. Gornik HL, Aronow HD, Goodney PP, et al. 2024 ACC/AHA/AACVPR/APMA/ABC/SCAI/SVM/SVN/SVS/SIR/VESS Guideline for the Management of Lower Extremity Peripheral Artery Disease: A Report of the American College of Cardiology/American Heart Association Joint Committee on Clinical Practice Guidelines. Circulation 2024;**149**:e1313–e1410. doi: 10.1161/CIR.0000000000001251

9. Mazzolai L, Teixido-Tura G, Lanzi S, et al. 2024 ESC Guidelines for the management of peripheral arterial and aortic diseases. Eur Heart J 2024. doi: 10.1093/eurheartj/ehae179

10. Internal Revenue Service, Department of the Treasury; Employee Benefits Security Administration, Department of Labor; Centers for Medicare & Medicaid Services, Department of Health and Human Services. Coverage of Certain Preventive Services Under the Affordable Care Act. https://www.federalregister.gov/documents/2015/07/14/2015-17076/coverage-of-certain-preventive-services-under-the-affordable-care-act (December 21, 2023)

11. Bonaca MP, Hamburg NM, Creager MA. Contemporary Medical Management of Peripheral Artery Disease. Circ Res 2021;128:1868–1884. doi: 10.1161/CIRCRESAHA.121.318258

12. Carmelli D, Fabsitz RR, Swan GE, et al. Contribution of genetic and environmental influences to ankle-brachial blood pressure index in the NHLBI Twin Study. National Heart, Lung, and Blood Institute. Am J Epidemiol 2000;151:452–458. doi: 10.1093/oxfordjournals.aje.a010230

13. Murabito JM, Guo CY, Fox CS, D’Agostino RB. Heritability of the ankle-brachial index: the Framingham Offspring study. Am J Epidemiol 2006;164:963–968. doi: 10.1093/aje/kwj295

14. Wang F, Ghanzouri I, Leeper NJ, Tsao PS, Ross EG. Development of a polygenic risk score to improve detection of peripheral artery disease. Vasc Med 2022;27:219–227. doi: 10.1177/1358863X211067564

15. Prive F, Aschard H, Carmi S, et al. Portability of 245 polygenic scores when derived from the UK Biobank and applied to 9 ancestry groups from the same cohort. Am J Hum Genet 2022;109:12–23. doi: 10.1016/j.ajhg.2021.11.008

16. Zhu K, Qian F, Lu Q, et al. Modifiable Lifestyle Factors, Genetic Risk, and Incident Peripheral Artery Disease Among Individuals With Type 2 Diabetes: A Prospective Study. Diabetes Care 2024. doi: 10.2337/dc23-1503

17. Bycroft C, Freeman C, Petkova D, et al. The UK Biobank resource with deep phenotyping and genomic data. Nature 2018;562:203–209. doi: 10.1038/s41586-018-0579-z

18. Koyama S, Wang Y, Paruchuri K, et al. Decoding Genetics, Ancestry, and Geospatial Context for Precision Health. medRxiv 2023. doi: 10.1101/2023.10.24.23297096

19. All of Us Research Program I, Denny JC, Rutter JL, et al. The "All of Us" Research Program. N Engl J Med 2019;381:668–676. doi: 10.1056/NEJMsr1809937

20. Genomes Project C, Auton A, Brooks LD, et al. A global reference for human genetic variation. Nature 2015;526:68–74. doi: 10.1038/nature15393

21. Boutin NT, Schecter SB, Perez EF, et al. The Evolution of a Large Biobank at Mass General Brigham. J Pers Med 2022;12. doi: 10.3390/jpm12081323

22. Aragam KG, Jiang T, Goel A, et al. Discovery and systematic characterization of risk variants and genes for coronary artery disease in over a million participants. Nat Genet 2022;54:1803–1815. doi: 10.1038/s41588-022-01233-6

23. All of Us All of Us Genomic Quality Report https://support.researchallofus.org/hc/en-us/articles/4617899955092-All-of-Us-Genomic-Quality-Report-

24. Yu S, Ma Y, Gronsbell J, et al. Enabling phenotypic big data with PheNorm. J Am Med Inform Assoc 2018;25:54–60. doi: 10.1093/jamia/ocx111

25. Castro VM, Gainer V, Wattanasin N, et al. The Mass General Brigham Biobank Portal: an i2b2-based data repository linking disparate and high-dimensional patient data to support multimodal analytics. J Am Med Inform Assoc 2022;29:643–651. doi: 10.1093/jamia/ocab264

26. Patel AP, Wang M, Ruan Y, et al. A multi-ancestry polygenic risk score improves risk prediction for coronary artery disease. Nat Med 2023;29:1793–1803. doi: 10.1038/s41591-023-02429-x

27. Ruan Y, Lin YF, Feng YA, et al. Improving polygenic prediction in ancestrally diverse populations. Nat Genet 2022;54:573–580. doi: 10.1038/s41588-022-01054-7

28. Patel AP, Khera AV. Advances and Applications of Polygenic Scores for Coronary Artery Disease. Annu Rev Med 2023;74:141–154. doi: 10.1146/annurev-med-042921-112629

29. Prive F, Arbel J, Vilhjalmsson BJ. LDpred2: better, faster, stronger. Bioinformatics 2021;36:5424–5431. doi: 10.1093/bioinformatics/btaa1029

30. Lambert SA, Gil L, Jupp S, et al. The Polygenic Score Catalog as an open database for reproducibility and systematic evaluation. Nat Genet 2021;53:420–425. doi: 10.1038/s41588-021-00783-5

31. Matsushita K, Sang Y, Ning H, et al. Lifetime Risk of Lower-Extremity Peripheral Artery Disease Defined by Ankle-Brachial Index in the United States. J Am Heart Assoc 2019;8:e012177. doi: 10.1161/JAHA.119.012177

32. Fahed AC, Wang M, Patel AP, et al. Association of the Interaction Between Familial Hypercholesterolemia Variants and Adherence to a Healthy Lifestyle With Risk of Coronary Artery Disease. JAMA Netw Open 2022;5:e222687. doi: 10.1001/jamanetworkopen.2022.2687

33. Klarin D, Tsao PS, Damrauer SM. Genetic Determinants of Peripheral Artery Disease. Circ Res 2021;128:1805–1817. doi: 10.1161/CIRCRESAHA.121.318327

34. van Zuydam NR, Stiby A, Abdalla M, et al. Genome-Wide Association Study of Peripheral Artery Disease. Circ Genom Precis Med 2021;14:e002862. doi: 10.1161/CIRCGEN.119.002862

35. Criqui MH, Matsushita K, Aboyans V, et al. Lower Extremity Peripheral Artery Disease: Contemporary Epidemiology, Management Gaps, and Future Directions: A Scientific Statement From the American Heart Association. Circulation 2021;144:e171–e191. doi: 10.1161/CIR.0000000000001005

36. van Rheenen W, Peyrot WJ, Schork AJ, Lee SH, Wray NR. Genetic correlations of polygenic disease traits: from theory to practice. Nat Rev Genet 2019;20:567–581. doi: 10.1038/s41576-019-0137-z

37. Truong B, Hull LE, Ruan Y, et al. Integrative polygenic risk score improves the prediction accuracy of complex traits and diseases. Cell Genom 2024;4:100523. doi: 10.1016/j.xgen.2024.100523

38. Natarajan P, Young R, Stitziel NO, et al. Polygenic Risk Score Identifies Subgroup With Higher Burden of Atherosclerosis and Greater Relative Benefit From Statin Therapy in the Primary Prevention Setting. Circulation 2017;135:2091–2101. doi: 10.1161/CIRCULATIONAHA.116.024436

39. Anand SS, Caron F, Eikelboom JW, et al. Major Adverse Limb Events and Mortality in Patients With Peripheral Artery Disease: The COMPASS Trial. J Am Coll Cardiol 2018;71:2306–2315. doi: 10.1016/j.jacc.2018.03.008

40. Kullo IJ, Jouni H, Austin EE, et al. Incorporating a Genetic Risk Score Into Coronary Heart Disease Risk Estimates: Effect on Low-Density Lipoprotein Cholesterol Levels (the MI-GENES Clinical Trial). Circulation 2016;133:1181–1188. doi: 10.1161/CIRCULATIONAHA.115.020109

41. Muse ED, Chen SF, Liu S, et al. Impact of polygenic risk communication: an observational mobile application-based coronary artery disease study. NPJ Digit Med 2022;5:30. doi: 10.1038/s41746-022-00578-w

42. Khera AV, Emdin CA, Drake I, et al. Genetic Risk, Adherence to a Healthy Lifestyle, and Coronary Disease. N Engl J Med 2016;375:2349–2358. doi: 10.1056/NEJMoa1605086

